# Specificity and Performance of Nucleocapsid and Spike-based SARS-CoV-2 Serologic Assays

**DOI:** 10.1101/2020.08.05.20168476

**Authors:** Zahra Rikhtegaran Tehrani, Saman Saadat, Ebtehal Saleh, Xin Ouyang, Niel Constantine, Anthony L. DeVico, Anthony D Harris, George K. Lewis, Shyam Kottilil, Mohammad M. Sajadi

## Abstract

There is an urgent need for an accurate antibody test for severe acute respiratory syndrome coronavirus 2 (SARS-CoV-2). In this paper, we have developed 3 ELISA methods, trimer spike IgA, trimer spike IgG, and nucleocapsid IgG, for detecting anti-SARS-CoV-2 antibodies. We evaluated their performance in comparison with four commercial ELISAs, EDI™ Novel Coronavirus COVID-19 ELISA IgG and IgM, Euroimmun Anti-SARS-CoV-2 ELISA IgG and IgA, and one lateral flow assay, DPP® COVID-19 IgM/IgG System (Chembio). Both sensitivity and specificity were evaluated and the causes of false-positive reactions were determined.

The assays were compared using 300 pre-epidemic samples and 100 PCR-confirmed COVID-19 samples. The sensitivities and specificities of the assays were as follows: 90%/100% (in-house trimer spike IgA), 90%/99.3% (in-house trimer spike IgG), 89%/98.3% (in-house nucleocapsid IgG), 73.7%/100% (EDI nucleocapsid IgM), 84.5%/95.1% (EDI nucleocapsid IgG), 95%/93.7% (Euroimmun S1 IgA), 82.8%/99.7% (Euroimmun S1 IgG), 82.0%/91.7% (Chembio nucleocapsid IgM), 92%/93.3% (Chembio nucleocapsid IgG).

The presumed causes of positive signals from pre-epidemic samples in commercial and in-house assays were mixed. In some cases, positivity varied with assay repetition. In other cases, reactivity was abrogated by competitive inhibition (spiking the sample with analyte prior to performing the assay). In other cases, reactivity was consistently detected but not abrogated by analyte spiking.

Overall, there was wide variability in assay performance using our samples, with in-house tests exhibiting the highest combined sensitivity and specificity. The causes of “false positivity” in pre-epidemic samples may be due to plasma antibodies apparently reacting with the analyte, or spurious reactivity may be directed against non-specific components in the assay system. Identification of these targets will be essential to improving assay performance.

## Introduction

There is an urgent need for an accurate serologic test for severe acute respiratory syndrome coronavirus 2 (SARS-CoV-2). Both antigen and antibody detection methods play important roles in disease management. Due to the high specificity of the rRT-PCR for detecting the presence of the virus during the acute phase, it is considered the gold standard for COVID-19 testing. Equally important, antibody-based tests can be used to identify infected individuals even when they are minimally symptomatic or asymptomatic or after symptoms are no longer present. Antibody-based tests are also key tools in population-level surveillance and determining protection against re-infection. In addition, the titers of specific antibodies against the SARS-CoV-2 Spike protein can be correlated to the neutralizing antibody levels in recovered people, and therefore can be potentially used for screening eligible convalescent plasma donors. At the time of writing this paper, the U.S. Food and Drug Administration (FDA) has authorized 158 molecular, 33antibody, and 2 antigen test under Emergency Use Authorization (EUA) (1). A number of these tests have been recalled due to poor performance.

It is known that the antibody levels in people recovered from infection to the closely related coronaviruses, such as SARS-CoV-1 and MERS-CoV, lasts months to years but seem to be non-durable (2-4). Although long-term information regarding the antibody response against SARS-CoV-2 is currently unavailable, we know the antibody response against SARS-CoV-2 can last at least several months. Therefore, the serologic tests evaluating specific antibodies against SARS-CoV-2 can be applied to show an immune response against this virus.

Despite their importance in disease management, the performance of many commercially available SARS-CoV-2 serologic tests have not been fully evaluated with large panels of samples, thus, their utility is questionable.

Because seasonal coronaviruses have conceivably infected the majority of the human population, cross reactivity to the common coronaviruses is an important concern in developing SARS-CoV-2 serology tests. These tests include ELISA, and chemiluminescent microparticle immunoassay (CMIA) or lateral flow immunoassays (as point of care tests), which target specific antibodies against viral spike or nucleocapsid proteins. We have developed 3 ELISA tests for detecting anti-SARS-CoV-2 antibodies and evaluated their performance characteristics in comparison to four commercial ELISA and one lateral flow assays.

## Methods

### Patient samples

A total of 100 plasma samples collected from PCR-confirmed COVID-19 patients and were used as the positive group. The samples were from patients hospitalized at the University of Maryland Medical Center with a diagnosis of COVID-19 in April-May 2020. Where possible, the last available sampling time point was used. In addition, a total of 300 pre-epidemic plasma/sera samples that were collected before the COVID-19 epidemic (2005-2019) served as negative controls. A summary of the samples used in this study are noted in Table 1. Samples were obtained from protocols approved by the University of Maryland, Baltimore IRB.

**Table 1.** Patient samples used in this study

|  |  |  |
| --- | --- | --- |
| Positive Samples | COVID-19, PCR confirmed | 100 |
| Pre-epidemic samples<br>(2005-2019) | HIV infected | 66 |
|  | HIV vaccinated | 70 |
|  | Blood donors | 164 |
| Total |  | 400 |

### In-house ELISAs

We developed three different ELISAs for the qualitative evaluation of the human serum or plasma for detection of IgG or IgA antibodies in human serum or plasma, targeting the SARS-CoV-2 trimer spike protein (IgG and IgA ELISA) and IgG antibodies targeting the SARS-CoV-2 nucleocapsid antigen (IgG ELISA).

#### A. Recombinant Proteins

SARS-CoV-2 trimer spike prefusion and trimerization-stabilized ectodomain (Cat. #46328, LakePharma, Hopkinton, MA) were used for developing the trimer spike IgG and IgA ELISAs. For the nucleocapsid IgG ELISA, the SARS-CoV-2 nucleocapsid protein was produced. Briefly, the nucleic acid sequence of the SARS-CoV-2 nucleocapsid (residues 1-419; isolate Wuhan-Hu-1; GenBank MN908947.3) was synthesized and cloned into OriGene expression vector by BlueHeron (Bothell, WA) along with C-terminal His tag. The plasmid was expressed in FreeStyle 293 cells and the transfected cells were harvested on day 7 post transfection. The recombinant nucleocapsid was affinity purified from the cell lysate by Ni-NTA Agarose (Qiagen) under native conditions. The purity of the purified nucleocapsid was evaluated by SDS-PAGE.

#### B. Developing and performing the in house ELISAs

The in-house ELISAs were performed by coating Immulon 2 HB 96-well flat bottom plates (ImmunoChemistry Technologies, Bloomington, MN) with 0.1 μg/well of the trimer spike or nucleocapsid proteins in TBS overnight at 4°C. Following blocking with Blotto (10% dried milk in TBS and 0.1% NP-40), the 1:100 diluted samples in Blotto was added and incubated for 1 hour at 37°C. After washing the plates with TBS, the antigen specific antibodies were incubated with alkaline phosphatase labeled goat anti human IgG or IgA antibodies (Southern Biotech, Birmingham, AL) for 1 hour at 37°C. After washing the wells, the BluePhos® Microwell (Seracare, Milford, MA) was added to each well as Substrate for 15 min at 37°C. The enzymatic reaction was stopped by adding KPL APstop™ Solution (Seracare, Milford, MA) and the signal was read at 650 nm.

### Commercial Assays

#### A. ELISAs

**The EDI™ Novel Coronavirus COVID-19 ELISA IgG** (Cat. # 1032, Epitope Diagnostics, Inc. San Diego, CA) was performed according to the manufacturer’s instruction. The assay is an ELISA that detects specific IgG antibodies in human serum or plasma by binding SARS-CoV-2 nucleocapsid on the plates. The cut-off value for negative and positive results was calculated by adding the calculated average of negative controls to 0.18 and multiplying to 0.9 and 1.1, respectively.

**The EDI™ Novel Coronavirus COVID-19 ELISA IgM** (Cat. #1033, Epitope Diagnostics, Inc. San Diego, CA) was performed according to the manufacturer’s instruction. The assay is based on the capture of total IgM antibodies in human serum or plasma and then detects antibodies binding SARS-CoV-2 nucleocapsid. The cut-off value for negative and positive results was calculated by adding the calculated average of negative controls to 0.1 and multiplying by 0.9 and 1.1, respectively.

**Euroimmun Anti-SARS-CoV-2 ELISA IgG and IgA** (Euroimmune, Germany; Cat # EI 2606-9601 G and EI 2606-9601 A, respectively) were performed according to the manufacturer’s instructions. The Euroimmun Anti-SARS-CoV-2 IgG and IgA tests are separate ELISAs that detect antibodies against the S1 subunit of the SARS-CoV-2 spike protein. Results were determined as a ratio of the signal of the samples to the average signal of Calibrators. The interpretation of the calculated ratios was performed as manufacturer’s recommendation. A ratio < 0.8 was considered as not elevated (Negative), ≥0.8 to <1.1 as indeterminate (borderline) and ≥1.1 as elevated (Positive).

#### B. Lateral Flow Assay

The DPP® COVID-19 IgM/IgG System (Chembio Diagnostics Systems Inc. Medford, NY) is a Dual Path Platform (DPP) system for qualitative rapid detection and differentiation of IgM and IgG antibodies to SARS-CoV-2 nucleoprotein in capillary “fingerstick” whole blood, venous whole blood, serum, and plasma samples. The test was performed at room temperature according to the manufacturer’s instruction. The results were read using the DPP Micro Reader that provides an objective result (positive and negative)

### Data Analysis

#### A) Cutoff calculation for in-house ELISAs

The cutoff values for the in-house ELISAs were determined by two different approaches. The first cutoff value was determined using the average OD plus 3 standard deviation (SD) of all 300 pre-epidemic samples. To determine the second cutoff value a ROC curve was generated for each assay using SPSS Statistics for Windows, version 16.0 (SPSS Inc., Chicago, Ill., USA). The area under curve (AUC) and the sensitivity and specificity at all the cutoff points were also calculated. Then, the optimal cutoff point was selected based on the point with the highest Youden index J (J = sensitivity + specificity - 1); this is the point on the curve in which the distance to diagonal line (line of equality) is maximum (5).

For each approach, the samples with ODs greater than the determined cutoff values were considered as positive and the samples with ODs equal or less than the cutoff values were considered as negative. The optimal cutoff value for each assay was determined based on the cutoff with the highest percent overall agreement (POA), which defined as the percentage of true positive and negative results divided to the total number of the samples (6). For calculation of the sensitivity and specificity of the commercial ELISAs the borderline results were excluded from calculation.

#### B) Calculation of the performance characteristics of all assays

Sensitivity and specificity were defined as the proportion of samples correctly identified as infected or non-infected based on their true status as determined by PCR for positives and the pre-epidemic samples for negatives. To determine and compare the performance of the tests, the AUC (for ELISA Tests) and the percent overall (clinical) agreement (for all assays) were calculated. In our study, all of the pre-epidemic samples are considered clinically negative and all of the PCR confirmed samples considered clinically positive, thus the percent overall agreement is equal to the overall clinical agreement.

### Soluble Antigen Competition Experiments to Determine Specificity of Presumptive False Positive Reactions

Antigen specificity of presumptive false positive reactions was assessed by using soluble SARS-CoV-2 trimer spike or nucleocapsid antigens to determine whether they inhibit reactivity of the specimen with the corresponding plate-bound antigen. For each assay the dilution of the starting plasma and concentration of the antigens used for the competition assay necessary to convert a true positive sample negative (or at least a 3-fold reduction) were determined. Depending on the assay, concentration of antigen used were 100-500ug/ml. Soluble trimer spike or nucleocapsid was pre-incubated with presumable false positive samples (direct plasma for spike and 1:100 plasma for nucleocapsid) for two hours at room temperature with shaking (Fisher Scientific isotemp, 125 RPM). To compensate for dilution of the specimens by pre-incubation, control specimens were pre-incubated with 100ug of bovine serum albumin (BSA) in PBS for comparison. After pre-incubation, the plates were evaluated by the various assays as described above.

## Results

### Determination of the optimal cutoff for in-house ELISAs

The cutoff values were calculated based on two different approaches and the optimal cutoff for each assay were determined by comparing the overall clinical agreement. Table 2 shows the performance of the in-house ELISAs. In the trimer spike IgG and IgA ELISA tests, the cutoff values calculated by ROC curve provided higher agreement with clinical status; thus, these values were selected for further evaluation of the assays. For the nucleocapsid ELISA the calculated cutoff values with the two approaches was partially equal (0.19 and 0.20).

**Table 2.**
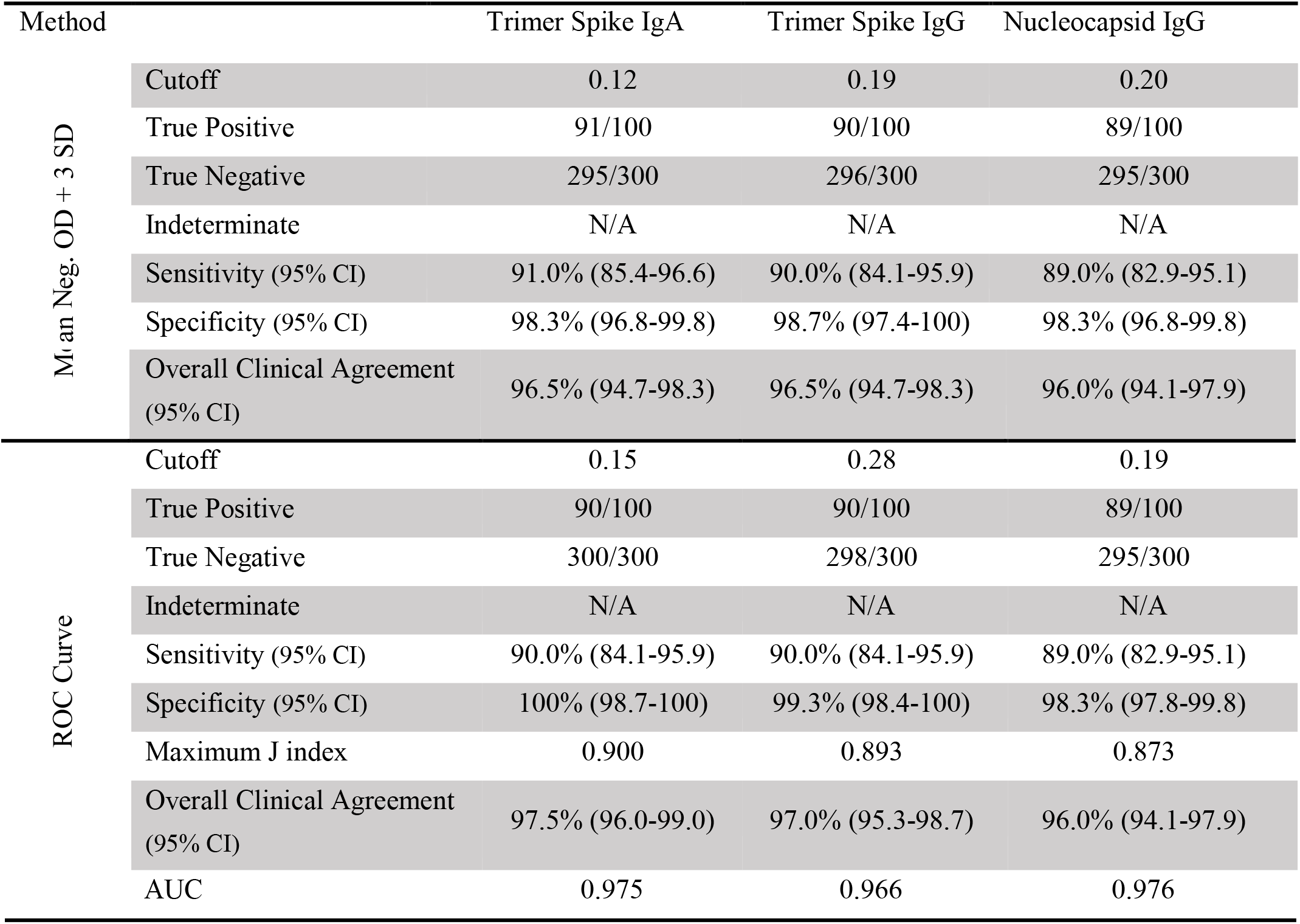
Performance of the in-house ELISAs using different calculated cutoff values

### Evaluation of the performance of all immunoassays

To determine the performance of the assays, the ROC curve analysis was performed for all ELISA assay. The obtained data are shown in Table 3 and Figure 1, and the supplementary file shows individual data from all experiments.

**Table 3.**
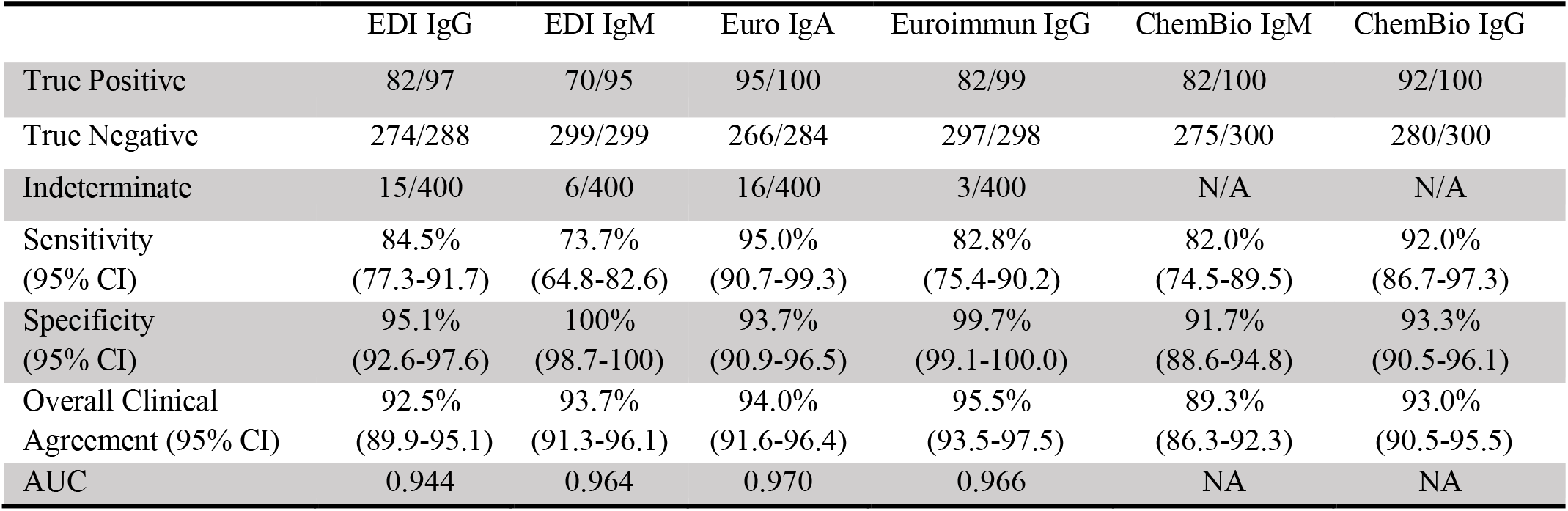
Performance of commercial assays

**Figure 1.**
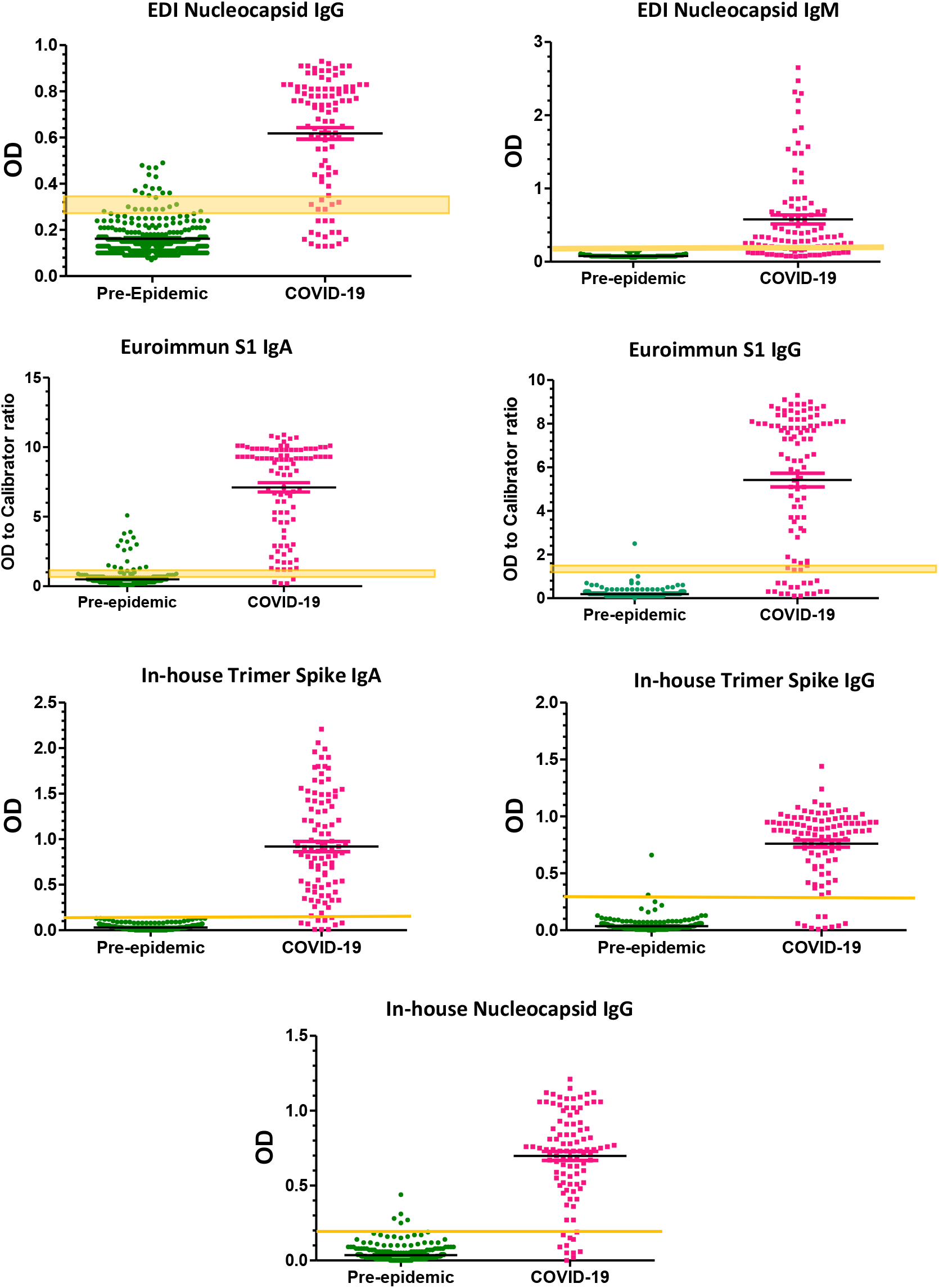
SARS-CoV-2 antibody assays data distribution. The data obtained for pre-epidemic and COVID-19 samples with all evaluated assays. The yellow lines show the cut-off values or ranges recommended by commercial assays or calculated cutoff values for in-house ELISAs. Black lines indicate median values with interquartile ranges.

### Soluble Antigen Competition Experiments

The in-house trimer spike IgA ELISA had no false positives and were not evaluated by soluble antigen competition. For the in-house trimer spike IgG ELISA, the one sample that was repeatedly false positive turned negative with spiking. For the in-house nucleocapsid IgG ELISA, four samples that were repeatedly false positive samples turned negative when nucleocapsid was added. The Eurommun SARS-CoV-2 IgG had one false positive that remained positive after spiking. Of the 18 false positive results of Euroimmun SARS-CoV-2 IgA, only one sample decreased substantially and turned to negative after adding the trimer spike protein. For the EDI SARS-CoV-2 IgG, 8 of 22 false positive and/or indeterminate pre-epidemic samples changed to negative when treated with the nucleocapsid protein. The false positive samples in the DPP rapid assay were rerun a number of times, with varying results obtained each time (almost half of the samples turned from positive to negative, or negative to positive on retesting). As such, they were excluded from further analysis in the spiking experiments. A summary of the spiking experiments can be found in Table 4 and the supplementary file has details of each experiment.

**Table 4.** Summary of the soluble antigen competition experiments

| Assay | Sample Status | Number of samples | Number (%) of samples that turned negative |
| --- | --- | --- | --- |
| In-house Trimer Spike IgG | False positive | 4* | 4 (100%) |
|  | Indeterminate | NA | - |
| In-house Nucleocapsid IgG | False positive | 1* | 1 (100%) |
|  | Indeterminate | NA | - |
| EDI IgG | False positive | 14 | 4 (29%) |
|  | Indeterminate | 8 | 4 (50%) |
| Euroimmun IgA | False positive | 18 | 2 (11%) |
|  | Indeterminate | 16 | 1 (6%) |
| Euroimmun IgG | False positive | 1 | 0 (0%) |
|  | Indeterminate | 2 | 1 (50%) |

\* One sample turned negative on retesting and not included
†8 out of 12 indeterminate pre-epidemic samples were tested in the Soluble Antigen Competition Experiments
NA= Not Applicable

## Discussion

Despite an urgent need and extensive efforts, the development of an efficient and fully validated serologic assay for detecting specific anti-SARS-CoV-2 antibodies are not yet available. In the present study, we developed and evaluated three ELISAs for detecting anti-trimer spike antibodies (IgG and IgA) and anti-nucleocapsid antibodies (IgG), and compared their performance with 5 commercial immunoassays, including 4 ELISAs (EDI™ Novel Coronavirus COVID-19 IgG and IgM, Euroimmun SARS-CoV-2 IgG and IgA) and one lateral flow immunoassay (DPP® COVID-19 IgM/IgG). For evaluation of the assays we used 100 COVID-19 PCR-confirmed samples from hospitalized patient and 300 pre-epidemic samples taken before the emergence of the disease in December 2019. The ratio of pre-epidemic to post-epidemic samples was 3:1 as we were particularly interested in studying the specificity of the assays and reasons for false positivity.

SARS-CoV-2 has four structural proteins (7-9), and among them the spike and the nucleocapsid are considered the main immunogens and are widely used in immunoassays. The nucleocapsid is a protein with a small size that can easily be produced and purified in prokaryotic or eukaryotic hosts in vast quantities. It has been shown that the anti-nucleocapsid antibodies appear earlier than the spike antibodies (10). Homology analysis show that the SARS-CoV-2 nucleocapsid has 90% amino acid identity to MERS-CoV and 28 to 49 percent identity to other alpha and beta coronaviruses (11). Therefore, applying the anti-nucleocapsid antibodies in ELISA tests may increase the clinical sensitivity of the assay if samples are drawn early. The same degree of similarity to other coronaviruses has also been seen in the spike protein, particularly the S2 domain (11). It has been suggested that using the S1 protein (and particularly the receptor binding domain) will improve the specificity of the immunoassays.

One area of concern is the potential for cross-reactivity with the four human seasonal coronaviruses. This concern can be addressed by designing a chimeric protein containing specific immunogenic regions of the spike and/or the nucleocapsid protein. However, it seems that until such a protein becomes available, it is still possible to provide high-efficiency immunoassays using these spike and nucleocapsid antigens. Here, we showed that the optimization of the assays with the minimum amount of coated antigen and conjugated detection antibodies along with the precise determination of accurate cutoff points improved the sensitivity and specificity of the in-house ELISAs.

Of the assays that employed spike protein (trimer for the in-house or subunit S1 for the Euroimmun), those with the best specificities include the in-house trimer spike IgA (specificity 100%), in house trimer spike IgG (99.3%), and Euroimmun IgG (99.7%), although the Euroimmun test also had close to an additional 1% indeterminate rate. The Euroimmun IgA performed poorly with a 93.7% specificity and 4% indeterminate results. The higher specificity of the Euroimmun IgG in comparison to the IgA test has been demonstrated in other studies (12, 13). Of the assays that employed nucleocapsid, those with the best specificities include the EDI-IgM (100% specificity) and the in-house nucleocapsid IgG (specificity 98.3%).

Some studies have shown different times to seroconversion to SARS-CoV-2 depending on the severity of the disease (14, 15), while those who are asymptomatic or minimally symptomatic may have lower or undetectable antibody levels (15, 16). Thus, both the timing of blood collections and severity of disease can potentially affect the sensitivity and specificity of the assays. We had three PCR positive samples, collected within the first week after the symptoms, which did not seroconvert during the time point of our study by any of the assays. It is possible that with longer follow-up time, some of these patients would have seroconverted. Use of a different patient population (truly convalescent samples) would likely increase the sensitivity in all the assays tested. Thus, the observed test indices must be viewed based on our population characteristics.

The overall clinical agreement is a useful parameter for evaluation and comparison of the performance characteristics of different assays. In the evaluated assays, the in-house ELISAs show higher clinical agreement than the commercial ones. The clinical agreement of the in-house trimer spike-IgA, trimer spike-IgG and nucleocapsid-IgG ELISAs was 97.5, 97.0 and 96.0%, respectively. While the highest agreement in the evaluated commercial assays belonged to the Euroimmun-IgG with 95.5% and the lowest one belonged to the Chembio-IgM with 89.3% agreement. It should be noted that, during the preparation of this paper, the FDA revoked the EUA for Chembio antibody test over concerns about accuracy of the test and high rate of false results (17). This suggests that the test was not verified to perform as expected.

Although it is expected that the presence of IgM antibody proves an earlier diagnosis for most infectious diseases, our findings suggest that the IgM antibodies are not detected substantially earlier than IgG antibodies. In our study, the two evaluated IgM assays, EDI-IgM and Chembio-IgM, showed the lowest sensitivity in comparison to the other assays. In addition, we did not have any IgM positive and IgG negative cases among the 100 COVID-19 samples, whereas the opposite was clearly noted (leading to a low sensitivity of the IgM assays). Similar findings have been shown in other SARS-CoV-2 and SARS-CoV-1 serodiagnostic assays (16, 18). However, we did not analyze serial samples, and thus no definite conclusions can be made as to the kinetics of the IgG and IgM responses.

In order to determine the cause for the false positivity in the various test formats, the false positive samples underwent competition experiments. In these experiments, the positive samples were pre-incubated with the analyte antigen, and thus, any significant cross-reactivity should result in a negative ELISA by competitive inhibition. For the in-house spike IgG and nucleocapsid IgG tests, all the samples that were repeatedly false positive became negative after the spiking, indicating that there was true cross-reactivity between the antibodies in the pre-epidemic samples and the SARS-CoV-2 spike or nucleocapsid antigens. Likewise, for the EDI nucleocapsid IgG, all the samples that were repeatedly false positive became negative after the spiking, indicating true cross-reactivity between the antibodies in the pre-epidemic samples and the SARS-CoV-2 spike and nucleocapsid antigens. This type of cross-reaction can be corrected by using the sample diluent buffers containing peptides of conserved nucleocapsid or spike (depending of the assay) of alpha and beta coronaviruses, or by designing a chimeric protein that contains highly specific sequences of antigens for coating.

By contrast, the competition experiment with the false positives from the Euroimmun SARS-CoV-2 IgG and Euroimmun SARS-CoV-2 IgA assays shows that most of these samples (18 of 19) are cross-reacting with something other than the coated S1 antigen used in the Euroimmun IgA. To reduce the number of false positive results in these assays, procedures such as re-optimization of the assay including revising the buffers, and increasing the purity of the coated antigen should be considered. Finally, about half the Chembio spiking experiments showed major problems with reproducibility, with the controls turning from positive to negative or vice-versa. Because of this, any attempt at interpretation was abandoned, and it should also be noted here that the FDA has revoked the EUA for this product.

In conclusion, we have evaluated the performance of several in-house ELISAs that incorporate the SARS-CoV-2 trimer spike and nucleocapsid antigens and compared them to the performance of the commercially available tests. All three in-house ELISA tests performed well, with high sensitivity and specificity, without the use of indeterminate results. Although the specificity of the best of these assays is high, even a specificity of 99% can be problematic in areas of low prevalence of SARS-CoV-2 infection i.e, positive predictive value. A high specificity of these assays is paramount because false positive results may lead to unnecessary quarantines, the closing of businesses, and concerns among family members and the community. It is essential to determine the cause of false positive results so that serologic assays can be improved. In addition, testing algorithms with more than one test may be necessary to rule out false positives by initial tests, such that has been the rule for HIV and hepatitis. Nevertheless, we have shown the potential of some assays to have specificities at, or approaching 100%, depending on their target isotypes, the antigens used, or on how the cutoff values are established.

## Data Availability

Data is available upon request.

## Acknowledgment

M.M.S. supported by NIH R01AI147870-01A1 and VA Merit Award IBX004525-01A2. A.H. supported by CDC U01CK000556-02-01

